# Urinary acetaminophen metabolites and clinical outcomes in premature infants

**DOI:** 10.1101/2024.05.29.24306893

**Authors:** Miguel Guardado, Dara Torgerson, Cheryl Chapin, Azuka Atum, Ryan D. Hernandez, B Ronald, Rebecca Simmons, Samuel Parry, Philip L. Ballard

**Author notes:** **Corresponding author:** Philip Ballard, Pediatrics, University of California, San Francisco, San Francisco CA 94194.

## Abstract

**BACKGROUND:** Extremely premature infants are treated with acetaminophen (APAP) for discomfort and patent ductus arteriosus. A recent study found an association between APAP metabolite levels in mothers’ breast milk and the diagnoses of both bronchopulmonary dysplasia (BPD) and retinopathy of prematurity (ROP) in their infants.

**METHODS:** Urine samples from 314 infants <29 weeks gestation in the TOLSURF and PROP studies were analyzed by untargeted UHPLC:MS/MS. We performed multivariate logistic regression and meta-analysis to examine associations between APAP metabolite levels and clinical outcomes.

**RESULTS:** 4-APAP sulfate was the highest detected and most abundant metabolite of 8 detected and was present in 98% of urines. In longitudinal studies (day 6-56), periods of elevated urinary 4-APAP-sulfate occurred in 24 of 28 infants and were of longer duration (10.1 vs 4.2 days, p=0.004) and higher levels (13.3 vs 5.6, p=0.013) in infants on enteral vs total parenteral nutrition. At both day 10 and 28 there were no significant associations between levels of APAP metabolites and BPD or ROP in all infants or only those on TPN or enteral feeds.

**CONCLUSION:** In two cohorts of premature infants, APAP metabolites were detected uniformly and levels were not associated with increased risk for two adverse clinical outcomes.

**Impact Statement:**

- Premature infants are treated with acetaminophen (APAP) for analgesia and closure of patent ductus arteriosus, however an association has been reported between APAP levels in maternal milk and infant bronchopulmonary dysplasia (BPD) and retinopathy of prematurity (ROP).
- In an untargeted metabolomic study of 2 cohorts of premature infants, the major urinary APAP metabolite was detected in most urine samples of all infants, and there were intervals of elevated levels.
- Using both longitudinal and cross-sectional analyses, we found no association between APAP levels and either BPD or ROP.
- Although APAP is known to have toxic effects at high doses, our findings suggest that APAP exposure, at doses experienced by infants in these cohorts, does not increase the risk for two adverse outcomes in the neonatal period.

## INTRODUCTION

Acetaminophen (paracetamol, APAP), an inhibitor of prostaglandin synthetase, is the most used analgesic world-wide. In the Neonatal Intensive Care Unit (NICU), APAP is used as an opioid-sparing agent for infant pain and discomfort and is also a front-line drug for closure of the patent ductus arteriosus (PDA) along with indomethacin and ibuprofen ^1,2^. In adults, there is an apparent association between APAP dose and hypertension, and APAP over-dose is the major cause of liver failure in the U.S. ^3,4^. It is also reported that in utero exposure to APAP is associated with increased risk for childhood dysregulated gonadal development, behavioral disorders and, notably, asthma ^5–10^.

Concern has been raised regarding the safety of APAP treatment in the NICU, particularly for infants <28 weeks gestational age with developmentally immature lungs and oxidative stress ^11–13^. Although meta analyses of clinical trials have found no increased risk of bronchopulmonary dysplasia (BPD) in infants treated for PDA with APAP vs nonsteroidal anti-inflammatory drugs ^2^, there are observational reports toward increased incidence of BPD in infants (mostly >28 weeks) exposed to APAP as an analgesic ^14,15^. Using a metabolomic approach, Santoro et al. recently reported an association between APAP metabolite levels in mothers’ breast milk at postnatal days 14 and 28 and the diagnosis of both BPD and retinopathy of prematurity (ROP) in their premature infants ^16^.

Untargeted metabolomic analysis involves separation and identification of low molecular weight chemicals by ultrahigh performance liquid chromatography and mass spectrometry (UHPLC-MS/MS) to provide quantitative data on hundreds of metabolites in a biofluid. Previous studies have examined the metabolome in premature infants using tracheal aspirate/lavage fluid ^17–20^, blood ^21,22^ and urine ^23–25^, identifying putative biomarkers for BPD or metabolic responses to treatments, however there is little metabolome-wide information available for xenobiotic chemicals present in the premature infant.

In the current study we used untargeted metabolomic analysis of urine from 2 cohorts of premature infants to investigate associations of APAP metabolites with two clinical outcomes (BPD and ROP) that are diagnosed later in the clinical course; we also determined levels of these metabolites over time in a subset of infants to provide profiles of APAP exposure in the NICU. Our findings provide new information on APAP metabolism in the premature infant and describe different patterns of exposure, however no significant associations were observed between metabolite levels and the two neonatal disorders.

## METHODS

### Clinical Cohorts

TOLSURF (Trial of Late Surfactant, ClinicalTrials.gov, NCT01022580) was a blinded, randomized, sham-controlled trial performed between 2010-2013 at 25 US centers to assess effects of late surfactant treatments on respiratory outcome. The trial design, infant characteristics and effects of late surfactant treatment have been reported ^26^. A total of 511 extremely low gestational age (<28 weeks) infants who required mechanical ventilation at 7–14 days were enrolled and received either surfactant (calfactant/Infasurf) or sham installation every 1–3 days. Clinical information included selected medications, age at full enteral feeds transitioning from total enteral nutrition (TPN), and respiratory parameters. Urine samples were collected (1-4/week for up to 8 weeks) using cotton balls in the diaper. Incidence of BPD at 36 weeks postmenstrual age, which was defined as a requirement for supplemental oxygen or positive pressure using an oxygen/pressure reduction test, and stage 2 ROP did not differ between treated and control groups.

PROP (Prematurity Respiratory Outcomes Program, NCT01435187) was an observational study performed between 2010-2013 at 8 US centers and designed to collect clinical data and biospecimens from infants <29 weeks’ gestational age for analysis of factors related to respiratory outcomes. Clinical information including selected medications, age at full enteral feeds transitioning from TPN, and respiratory parameters were obtained as in the TOLSURF study. Urine samples were collected at approximately days 7, 14, 21 and 28 using cotton balls in the diaper. BPD was defined as for the TOLSURF study. A total of 835 infants <29 weeks’ gestational age were enrolled in PROP at 1-7 days with characteristics as previously described ^27^.

Plasma was obtained during 2015 from a cohort of 12 newborn premature infants (gestational age 30.6±4.2 weeks, 11 African American and 1 Non-Hispanic White) and their mothers at the time of uncomplicated premature birth at University of Pennsylvania hospital-affiliated obstetric practices.

The research protocols for all studies were approved by the Institutional Review Boards of the participating institutions, and a parent of each infant provided written informed consent.

### Metabolomic Analysis

Untargeted metabolomic analysis of relative levels of metabolites in urines and plasma by UHPLC-MS/MS was performed by Metabolon Inc (Morrisville, NC) as described ^22,24,25^. For cross-sectional analysis of chemical levels, metabolomic analyses were performed on a total of 644 urine samples from subsets of infants (171 TOLSURF infants and 143 PROP infants) who had urine samples available at two time points (day ∼10 and day ∼28). A longitudinal analysis of metabolite levels was performed in 302 urines from 28 TOLSURF infants with ≥5 (mean/sd 11±4) urine samples available over ≥19 (mean/sd 35±8) days; 25 of the 28 infants for the longitudinal study were also included in the cross-sectional study at 2 time points. In these analyses we evaluated all metabolite levels and separately those defined as elevated (>2-fold of median).

The metabolite dataset comprised a total of 1435 compounds of both known identity (named by Metabolon as of July 2022, 77%) and unnamed (23%) chemicals. Area under the peak was obtained for each biochemical and adjusted for any inter-assay (batch) variability and urine osmolality and then rescaled setting the median value of all samples for each chemical to 1. For analysis of levels of metabolites between samples, we used these median-normalized values.

Missing values (below the limit of detection), if any, were imputed using the minimum detectable value for each compound. To compare relative amounts of APAP metabolites in a urine sample, we used area under the peak values and normalized to the value for 4-APAP sulfate, the most abundant APAP metabolite, for each infant.

### Statistical Analysis

Univariate comparisons between outcomes and APAP metabolite abundance were analyzed using Fisher’s exact test (detected vs not detected) and the Mann-Whitney U-test (levels) to adjust for incomplete parametric transformation with rank normalization. We performed multivariate logistic regression to identify chemicals that varied in level by clinical outcome (BPD, ROP), adjusting for potential confounders of clinical importance: birthweight, sex, corticosteroid treatment, type of nutrition (TPN vs enteral nutrition) and maternal self-identified race/ethnicity. Logistic regression was performed in python 3.8 using the statsmodel package. Analyses were performed within each study cohort separately, and results combined in a meta-analysis using the metafor package in R 4.2.1 to account for study and experimental batch effects between the two cohorts (TOLSURF and PROP). All source code is available on GitHub. Metabolite data are presented as mean±sd or as median and interquartile (IQ) range.

## RESULTS

The gestational age for all infants with metabolomic data was mean/sd 25.3±1.2 weeks and birth weight was 727±170 g; details of clinical characteristics of these cohorts have been reported ^24,25^. Untargeted metabolomic analysis identified 233 named xenobiotic chemicals, including 9 APAP-related chemicals.

### Detection and Relative Abundance of APAP Metabolites

Three metabolizing pathways for APAP (sulfation, glucuronidation and oxidation (via CYP2e1) have been described. Our study detected unaltered APAP and a total of 8 APAP metabolites in infant urine samples: 4 sulfated metabolites, 2 glucuronide metabolites, and 2 cysteinyl metabolites (via oxidation). Detection rates ranged from 7.6 % (2-methoxyAPAP glucuronide) to 98.2% (4-APAP sulfate). 4-APAP sulfate was the most abundant urinary metabolite, based on relative values for area under the peak for each urine sample, with all other APAP metabolites at <20% (Table 1).

There were high correlations between levels of 4-APAP sulfate and the other metabolites by linear regression with r values ranging from 0.58 (2-hydroxyAPAP sulfate) to 0.90 (APAP) with all p values <0.0003. Figure 1 shows four examples of regression plots for 4-APAP sulfate vs other APAP metabolites in the same sample, illustrating high correlations between 4-APAP-sulfate and APAP (Fig. 1a), another sulfate derivative (Fig. 1b), a cysteinyl derivative (Fig 1c) and a glucuronide derivative (Fig 1d).

**Figure 1.** Comparison of abundance between selected urinary APAP metabolites. a, 4-APAP sulfate vs APAP; r = 0.79, p = 3.6e-28, n = 129. b, 4-APAP sulfate vs 3-(methylthio)APAP sulfate; r = 0.59, p = 2.9e-20, n = 203. c, 4-APAP sulfate vs 3-(N-acetyl-cystein-S-yl) APAP; r = 0.90, p = 7e-108, n = 296. d, 4-APAP sulfate vs 4-acetamidophenylglucuronide; r = 0.75, p = 5.1e-26, n = 140. Data are unadjusted area under the curve values from MS:MS for urines where both metabolites were detected and quantified in the sample. Analysis by linear regression with n values as listed. Note 4-fold difference between vertical and horizontal axis.

To assess exposure of infants to APAP before birth, we examined detection rate and relative abundance of metabolites in cord and maternal plasma samples from a separate cohort of premature infants using the same metabolomic platform for analysis (Table 1). The range of detection rates for APAP and 7 metabolites were similar for cord (33-75% of infants) and maternal samples (25-58% of mothers). The abundance of all APAP metabolites (relative to 4-APAP sulfate) was similar in cord and maternal plasma except for the two cysteinyl derivatives that were lower in fetal plasma. Compared to infant urine, detection rates for maternal plasma were significantly higher for 3 metabolites: 2-hydroxyAPAP sulfate (58% maternal vs 12% urine), 3-(cystein-S-yl)APAP (42% vs 17%) and 2-methoxyAPAP glucuronide (33% vs 8%), and lower for 2 metabolites: 4-APAP sulfate and 3-(N-acetyl-cystein-yl) APAP. There was a high relative abundance of 4-acetamidophenylglucuronide in maternal (0.90) and cord plasma (1.56) vs urine (0.02), representing metabolism in adults vs premature infants, which is consistent with increased glucuronidation during postnatal development ^28–30^.

### APAP Levels in Time Course Studies

For time course studies in a subset of 28 TOLSURF infants, we used 4-APAP sulfate as a representative urinary indicator of APAP exposure based on the high detection rate and strong correlations of levels with those of other derivatives. In serial collections of urine, most (23/28, 82%) TOLSURF infants had at least 1 urine sample with elevated levels of 4-APAP sulfate (defined as >2-fold of median). Time course plots (Fig. 2) show examples of periods of elevated urinary 4-APAP sulfate during total parenteral nutrition (TPN) (panel a) and while on full enteral nutrition (panels b and c) compared to an infant with levels <1.0 in all samples (panel d), illustrating the episodic nature of increased APAP levels imposed on as lower baseline concentration.

**Figure 2.** Time course for levels of urinary 4-APAP sulfate in 4 infants. a, 25.6-week gestation infant with BPD, ROP and PDA who remained on TPN until day 46 with 2 episodes of elevated metabolite. b, 25.1-week gestation infant with ROP who transitioned to full enteral feeds on day 14; 4-APAP sulfate was elevated at day 10 and between day 18 and 33 (note higher scale of x axis). c, 25.0-week gestation infant with BPD and ROP on full enteral feeds from day 28; 4-APAP sulfate was elevated from day 21 to day 48. d, 24.1-week gestation infant with BPD and ROP on full enteral feeds from day 24; all values are less than 1.

We examined APAP levels from samples during TPN vs enteral nutrition with breast milk as an exploratory approach for assessing the possible contribution of maternal APAP to the infants (Fig. 3a). The level of 4-APAP sulfate in samples during peaks were higher on full enteral nutrition (median 13.3, IQ 8.1/28.6) than during TPN (median 5.6, 3.8/10.3, p = 0.011 by Mann-Whitney test), and the duration of elevated levels was 10.1±7.5 days vs 4.2±2.0, p=0.004 for full enteral nutrition vs TPN, respectively. The percent of 4-APAP sulfate values >2 and the median level for all samples did not differ by feeding status.

**Figure 3.** Summary of time course parameters for elevated (>2-fold of the median value) for infants with time course data. a, 4-APAP sulfate levels in infants on TPN (n=17-26 infants) vs enteral nutrition (n=17-25 infants); the level and duration of 4-APAP sulfate peaks are ∼3-fold greater on enteral feeds (p≤0.01). b, summary of time course parameters for 4-APAP levels in infants with (n=17 infants) vs without BPD (n=11); there are no significant differences.

Comparing the same 4 time-course parameters between infants with (n=17) and without (n=11) BPD revealed no significant differences (Fig. 3b). Similarly, there were no differences detected examining ROP as the outcome (data not shown). Thus, based on the 98% detection rate of urinary 4-APAP sulfate, all infants were continually exposed to APAP throughout the first 2 postnatal months. Most infants had one or more periods of exposure to higher doses, and levels were higher and of longer duration while receiving enteral nutrition compared to TPN feeds; however, there were no significant associations with the two clinical outcomes in these longitudinal studies.

### APAP Metabolite Levels in Cross-sectional Analyses

Urinary levels of 4-APAP sulfate varied markedly between 314 infants at both day 10 and 28 (2,163- and 15,692-fold, respectively), which is consistent with intermittent exposure to higher doses of APAP during the first postnatal month. There was no difference by time point (day 10 vs 28) in number of infants with 4-APAP sulfate values elevated >2.0 (39.2 vs 37.0%), nor in median values of levels >2 (8.6 and 9.7, respectively). In day-28 samples from TOLSURF infants, feeding status (TPN n=61; full enteral nutrition n=110) did not significantly influence either the percentage of infants with elevated values (34.4 vs 40.0%, respectively) or median level for elevated values (8.8 vs 9.9, respectively).

Using multivariate logistic regression and meta-analysis of the two infant cohorts, we examined the cross-sectional metabolomic data at days 10 and 28 for associations of levels of APAP and all detected APAP metabolites with clinical outcomes. BPD, which was defined as a need for positive pressure or supplemental oxygen at 36 weeks, occurred in 56.7% of the 314 study infants, and ROP of stage ≥2 occurred in 53.9%. Analyses at day 28 also were performed separately for infants on TPN vs full enteral nutrition because of the marked effect of feeding status on the infant metabolome ^25^ and the reported associations between APAP levels in breast milk and premature infant outcomes ^16^.

In both day-10 and day-28 urine samples there were no significant associations (p<0.05) between detection rate or levels of APAP metabolites and BPD by univariate analyses (data not shown). In multivariate logistic regression of APAP metabolite levels, adjusting for birthweight, sex, corticosteroid treatment, type of nutrition (TPN vs enteral nutrition) and maternal self-identified race/ethnicity, no significant associations were found for all samples at day 10 (Table 2, data columns 1-6) or at day 28 (columns 7-12).

Previously, we reported that levels of APAP metabolites were not associated with feeding status using logistic regression that was adjusted for selected clinical variables, predicting that separate analysis for infants on TPN or on enteral nutrition would not show associations with BPD or ROP ^25^. To directly test this possibility, we also examined APAP associations with BPD and ROP by multivariate logistic regression in day-28 infants separately by nutritional source, not adjusting for feeding status: TPN = 188 infants; enteral nutrition = 126 infants. There were no significant associations for infants on TPN (Table 3, data columns 1-6) and one significant association by meta-analysis for infants on enteral nutrition (columns 7-12): 3-(methylthio)APAP sulfate (OR 0.91, CI 0.82/1.00, p=0.045, last column), indicating lower metabolite levels in infants with BPD. OR values for the other metabolites on enteral nutrition were all <1 with insignificant p values ranging from 0.07 to 0.76 in the cross-cohort meta-analysis. Six metabolites had p<0.05 and OR <1 in the TOLSURF cohort on enteral nutrition (columns 19, 20), consistent with decreased risk of BPD, but statistical significance was not observed for the PROP cohort. The same analyses were carried out for ROP, and there were no significant associations of APAP metabolites at either time point (Table 4) or by feeding status (Table 5). Thus, levels of APAP urinary metabolites were not associated with increased risk for two outcomes that are diagnosed later in the clinical course after exposure to APAP.

## DISCUSSION

In this study we used untargeted metabolomics to examine urinary metabolites of APAP and associations with clinical outcomes in extremely low gestation premature infants. The major metabolite 4-APAP sulfate was detected in 98% of samples collected between 6 and 59 days; most infants had intervals of elevated levels that were higher and of longer duration while on enteral feeds. At both 10 and 28 days, levels of APAP and eight APAP metabolites were not significantly associated with BPD or ROP, providing evidence derived from the infant urinary metabolome that APAP exposure is not associated with these adverse clinical outcomes.

As reviewed ^11,12^, there has been concern regarding the safety of APAP in infants based on animal data, individual pharmacokinetic data and results from some non-randomized clinical studies. In recent metabolomic studies with regression analyses at days 14 and 28, Santoro et al. ^16^ reported significant associations between levels of APAP metabolites in breast milk with BPD (p values 0.009-0.04 and OR values 3.1-6.7) and between APAP levels and ROP (p 0.04 and OR 2.7). Our study in infant urine does not confirm this association; in fact, most OR values were <1 including the one significant association identified for 3-(methylthio)APAP sulfate (meta-analysis p=0.05), suggesting possible protection by APAP.

Both the Santoro and our study were conducted in extremely low gestation newborns of similar gestational and birth weight (breast milk/urine mean gestational age 26.3/25.3 weeks and mean birth weight 840/727 g) who were born in the same era (breast milk 2009-2011, urine 2010-2013) and used the same metabolomics platform (Metabolon), which provided consistency for the analytic approach and metabolite library. Our definition of BPD (physiologic requirement for supplemental oxygen and/or positive pressure) was somewhat more rigorous than in the Santoro study, which may account for the somewhat lower incidence of BPD in our cohorts (TOLSURF 62.0%, PROP 50.3%) compared to Santoro (65.3%). The most important difference between the two studies was measurement of APAP in the infant urine vs in maternal breast milk, which may not necessarily reflect APAP levels in the infant because of variable milk consumption and different relative abundance of metabolites after maternal metabolism. The parallel APAP association results for BPD and ROP, in both the Santoro study and ours, is consistent with these disorders tending to occur in the same, sicker infants. We believe that our results provide strong evidence for lack of a clinically significant association between APAP exposure, at the levels experienced by TOLSURF and PROP infants, and both BPD and ROP. However, a definitive conclusion may require an additional study with measurements in both breast milk and in the infant.

Premature infants can be exposed to APAP by direct administration in the NICU and secondary to maternal intake and receiving breast milk. Because APAP is considered safe (up to 3 g/day) during pregnancy, many infants are exposed in utero intermittently throughout gestation, including during labor both preterm and at term. Our studies with cord plasma found 4-APAP sulfate in 75% of preterm infants at birth (and their mothers) with a similar detection rate in term infants (data not presented); this is consistent with the reported use of APAP in pregnancy ^31,32^. With a half-life of ∼4 hours in infants, the drug should be fully cleared after a few days, and thus prenatal exposure does not explain the uniform presence of APAP in infant urine collected at 7-14 days. In the UCSF NICU, APAP is routinely prescribed as an analgesic for infant discomfort on an as-needed basis at ∼10 mg/kg q 6 hours. Our findings of detectable urinary APAP in 142 of 143 samples from infants on TPN (up to 46 days postnatal age) are consistent with continual use for discomfort in this population of ill infants.

Following the report by Hammerman et al in 2011 ^33^, APAP has increasingly been used to promote closure of the PDA, initially as a back-up drug to indomethacin and ibuprofen and currently often as the front-line drug, and closure rate is serum level-dependent ^34^. APAP dosing information was not collected in TOLSURF or PROP, however it is likely that some of the infants enrolled between 2012 and 2013 received APAP for PDA closure (at doses of 10-20 mg/kg q 6 hours with adjustment of dose based on plasma levels in some units). The highest levels of 4-APAP sulfate were observed in infants (18 of 19) with samples collected while on enteral feeds, and elevated levels were episodic rather than continuous in most infants. Most of these samples were collected beyond the age when PDA is usually treated, and we conclude that these elevated levels reflect intermittent exposure of infants to APAP in breast milk from their mothers or from banked donor milk, which is consistent with the data of Santoro et al. ^16^. The recommended adult dosing at ∼15 mg/kg q8 hours is higher than the dose used in infants for analgesia (10 mg/kg q 6 h), and it is possible that some mothers of premature infants exceed the recommended dose of APAP, leading to higher levels transferred to infants via breast milk. Also, some APAP metabolites may be concentrated in maternal milk or have different clearance rates in the infant. Additional studies of APAP pharmacokinetics in extremely low gestation newborns are needed.

The hepatotoxicity associated with high APAP doses is well established and results from production by CYP2E1 of the metabolite N-acetyl-p-benzyl quinone; this toxic metabolite was not analyzed in our study, however we did find low levels of APAP cysteinyl derivatives that result from binding of N-acetyl-p-benzyl quinone to cysteine in proteins. Cysteinyl derivatives are also found in serum of most adults taking APAP at therapeutic doses at concentrations below the threshold associated with hepatotoxicity ^35^. CYP2E1 is also present in the lung and may contribute to the reported association between APAP and chronic obstructive lung disease and asthma ^8–10^. Because N-acetyl-p-benzyl quinone is inactivated by glutathione, toxic effects would presumably be greater under oxidative conditions with depletion of glutathione such as occurs in infants with lung disease who experience oxidative stress. This could lead to impaired alveolization in the developing lung and increased likelihood of BPD. However, evidence for a causal link between APAP and BPD remains lacking.

Strengths of our study include the relatively large number of infants studied, the multi-institutional cohorts, the detailed clinical data base and both cross-sectional and longitudinal analyses; however, there are limitations. Some aspects of clinical care varied between institutions, and information on dose level and schedule of APAP treatment was not collected. Because the use of APAP for ductal closure has increased since 2013, exposure to APAP currently is likely greater for many infants than in our trials. Although clinical data were not collected in TOLSURF for the type of milk (breast vs formula) at initiation of enteral feeds, we assume that most if not all infants received full or partial breast milk feeds. This was the standard of care in the NICU in 2010-2013 in compliance with the guidelines of the American Academy of Pediatrics ^36^. Our previous metabolomic analysis of urines from infants on TPN vs enteral nutrition also supports this conclusion: in day-28 samples, levels of 45 of 69 chemicals in the Food Component sub pathway of Xenobiotic chemicals were significantly (p<0.05) different by feeding status, and levels were higher for infants on enteral feeds for 91.1% of these metabolites. An example is cinnamoylglycine, the glycine derivative of cinnamic acid that is abundant in fruits and vegetables: mean level on TPN vs enteral feeds was 0.165, p=0.000017 by logistic regression ^25^. Finally, we cannot rule out the possibility that confounding variables that were not assessed contribute to the lack of observed associations of outcomes with APAP metabolites.

In summary, the major APAP metabolite was detected in most urine samples of all infants, and intervals of elevated levels occurred that were higher and of longer duration during enteral feeds. Using both longitudinal and cross-sectional analyses, we found no association in two infant cohorts between APAP levels and BPD or ROP. Although APAP is known to have toxic effects at high doses, our results suggest that APAP exposure, at doses experienced by infants in our cohorts, does not increase the risk for two adverse outcomes in the neonatal period. Importantly, this finding needs confirmation and does not address the likely increased risks associated with prolonged exposure to higher doses in premature infants.

## Abbreviations

BPD: bronchopulmonary dysplasia
ROP: retinopathy of prematurity
APAP: acetaminophen
UHPLC-MS/MS: ultrahigh performance liquid chromatography-mass spectrometry
TPN: total parenteral nutrition

## Data availability

The datasets generated and analyzed during the current study are available from the corresponding author upon reasonable request.

## Patient ID Use

Infant ID’s are randomized and only accessible to the members of the research group involved in this study.

## Deletions

ROP of stage ≥2 occurred in 285 (55.8%) of TOLSURF infants and was more prevalent in infants with BPD (74% vs 61.5%, p = 0.0025 by chi-squared test).

BPD the treated (61.8%) and control (59.6%) groups.

The median detection rate for all metabolites was 93.6% (IQ 50.3/100).

